# Who is leading medical AI? A systematic review and scientometric analysis of chest x-ray research

**DOI:** 10.64898/2026.04.02.26349884

**Authors:** Constanza Vásquez-Venegas, Api Chewcharat, Richard Kimera, Nicholas Kurtzman, Marianna Leite, Naira Link Woite, Ishvica J. Muppidi, Rachel J. Muppidi, Xioli Liu, Erika P. Ong, Ridhima Pal, Christopher Myers, Sebastian Salzman, Jonathan Sønderbo Patscheider, Taniya Raji John, Mwavu Rogers, Mathew Samuel, Juan Luis Santana-Guerrero, Shahar Yaacob, Rodrigo Gameiro, Leo A. Celi

## Abstract

Computer vision models for chest X-ray interpretation hold significant promise for global healthcare, but their clinical value depends on equitable development across diverse populations. We conducted a scientometric analysis to examine authorship patterns, geographic distribution, and dataset origins to assess potential disparities that could affect clinical applicability.

We systematically reviewed literature on computer vision applications for chest X-rays published between 2017-2025 across multiple databases, including PubMed, Embase and SciELO databases. Using Dimensions API and manual extraction, we analyzed 928 eligible studies, examining first and senior author affiliations, institutional contributions, dataset provenance, and collaboration patterns across different income classifications based on World Bank categories.

High-income countries dominated research leadership, representing 55.6% of first authors and 59.7% of senior authors; no first authors were affiliated with low-income countries.

China (16.93%) and the United States (16.72%) led in first authorship positions. Most datasets (73.6%) originated from high-income settings, with the United States being the largest contributor (40.45%). Private datasets were most frequently used (20.52%). Cross-income collaborations were rare, with only 3.9% of publications involving partnerships between high-income and lower-middle-income countries.

Findings reveal substantial disparities in who shapes computer vision research on chest X-rays and which populations are represented in training data. These imbalances risk developing AI systems that perform inconsistently across diverse healthcare settings, potentially exacerbating healthcare inequities. Addressing these disparities requires coordinated efforts to develop globally representative datasets, establish equitable international collaborations, and implement policies that promote inclusive research practices.

**Author Summary:** In this study, we examined the global landscape of research involving computer vision applied to chest X-rays. While these technologies have the potential to significantly improve healthcare worldwide, their effectiveness depends on being developed and tested using data from diverse populations. We analyzed nearly one thousand scientific articles and found that research leadership and data sources are heavily concentrated in high-income countries, particularly the United States and China.

Our findings reveal a concerning gap because people in low-income regions, who often face the highest burden of respiratory diseases, are almost entirely absent from the research process as both authors and data contributors. This imbalance creates a risk that medical artificial intelligence may not perform reliably when used in different parts of the world, which could accidentally worsen existing health inequalities. We argue that the scientific community must prioritize international partnerships that treat researchers from developing nations as equal leaders. By making medical data more diverse and accessible, we can ensure that these powerful diagnostic tools benefit patients everywhere, regardless of their location or economic status.

## Introduction

Computer vision (CV) applications in healthcare represent one of the most promising frontiers in medical artificial intelligence and have the potential to transform clinical workflows worldwide. These technologies leverage deep learning architectures to extract diagnostic insights from visual data, including X-rays, computed tomography (CT) scans, and other medical imaging modalities.^1,2^ In particular, CV application to chest X-rays (CXRs) has attracted substantial research attention due to CXRs’ central role in diagnosing and monitoring respiratory conditions, which remain among the leading causes of global morbidity and mortality.^3^

The clinical appeal of automated CXR interpretation stems from multiple factors: the high volume of CXRs performed globally, persistent shortages of radiologists in many regions, and the potential for computational approaches to detect subtle patterns that might escape human observation.^4–6^ Recent studies have demonstrated that CV algorithms can achieve radiologist-level performance in detecting conditions ranging from tuberculosis to COVID-19.^7–9^

However, the clinical value of these technologies critically depends on their ability to perform reliably across diverse patient populations and healthcare settings. Emerging evidence suggests that many AI systems exhibit substantial performance disparities when deployed outside their development environment.^10–13^ This “generalizability gap” stems largely from dataset biases—models trained predominantly on data from specific demographic groups, healthcare systems, or geographic regions often underperform when applied to underrepresented populations.^14^ These disparities are particularly concerning given the global distribution of respiratory disease burden. Lower-income and middle-income countries (LMICs) experience disproportionately high rates of tuberculosis, pneumonia, and other respiratory conditions that could benefit from automated diagnostic support; nevertheless, these same regions may be underrepresented in the datasets that train CV systems.^3,15,16^

Therefore, understanding the global landscape of CV research on CXRs is crucial for identifying potential blind spots in the field. Scientometric analysis provides a systematic approach to mapping knowledge production networks, revealing who contributes to research, where datasets originate, and how collaboration patterns influence technological development.^17,18^ By applying these methods to CV research on CXRs, we can evaluate whether the current research ecosystem promotes or hinders the development of globally equitable diagnostic technologies.

This study presents a comprehensive scientometric analysis of the literature on CV applications to CXRs published between 2017 and 2022. We examined authorship patterns, institutional affiliations, geographical distribution, and dataset provenance to quantify disparities in research contribution and representation. Our analysis addresses three critical questions:

1. How equitably is research leadership distributed across world regions and income groups?
2. What are the predominant patterns of international collaboration in this field?
3. How representative are the datasets used to develop and validate CV models for CXR interpretation? By identifying structural imbalances in the research landscape, this work aims to inform more inclusive approaches to developing medical AI that can benefit diverse global populations.

## Results

### I. Search Strategy

A systematic search (Figure 1) was conducted in the PubMed, Embase, and SciELO databases, yielding 1555 articles. After removing 3 duplicate records, 1552 unique articles remained for screening. Titles and abstracts were assessed, leading to the exclusion of 455 articles. Subsequently, a full-text review was performed, resulting in the exclusion of an additional 113 articles. In total, 984 studies were included in the qualitative synthesis. For the quantitative analysis, 928 studies were retained following an evaluation of quality considerations.

**Fig. 1.**
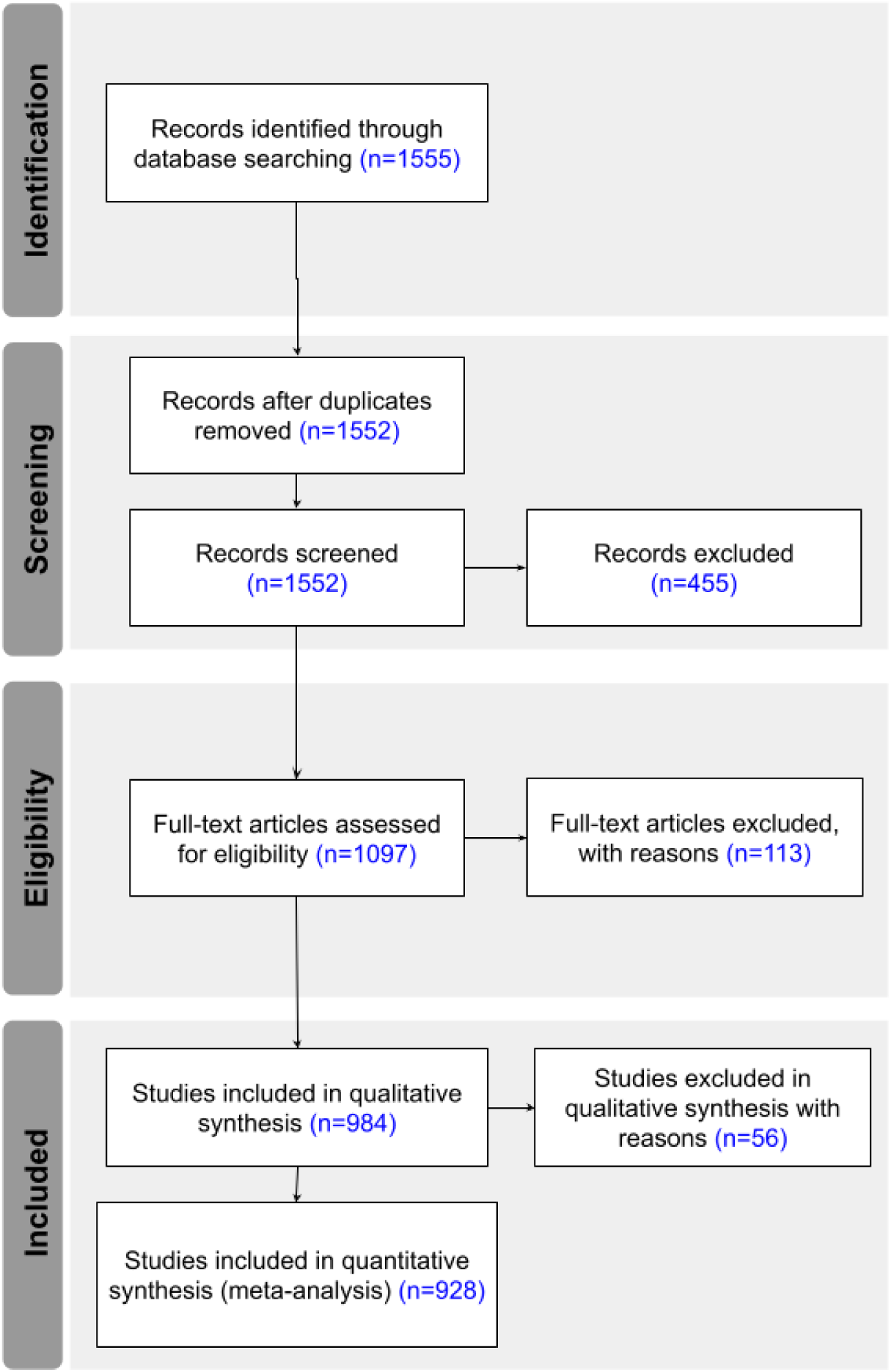
Flow diagram for article inclusion and exclusion based on PRISMA methodology.

### II. Scientometric Analysis

To evaluate the global landscape of CV research applied to CXRs, we conducted a scientometric analysis focusing on key aspects of knowledge production and dissemination. This section presents findings on publication trends, leading institutions, and the geographical distribution of research contributions. We assess the dominance of high-income institutions, analyze collaboration patterns between countries of different income levels, and examine the representation of datasets across regions.

The journal with the highest number of publications on CV applied to CXR was *Scientific Reports*, with 93 publications (10.05%), followed by *PLOS One* (4.11%) and *Computers in Biology and Medicine* (4.00%). Notably, from a total of 239 journals included, no single journal accounts for a dominant share of the literature, suggesting a broad but somewhat fragmented distribution of research outputs (Figure 2).

**Fig. 2.**
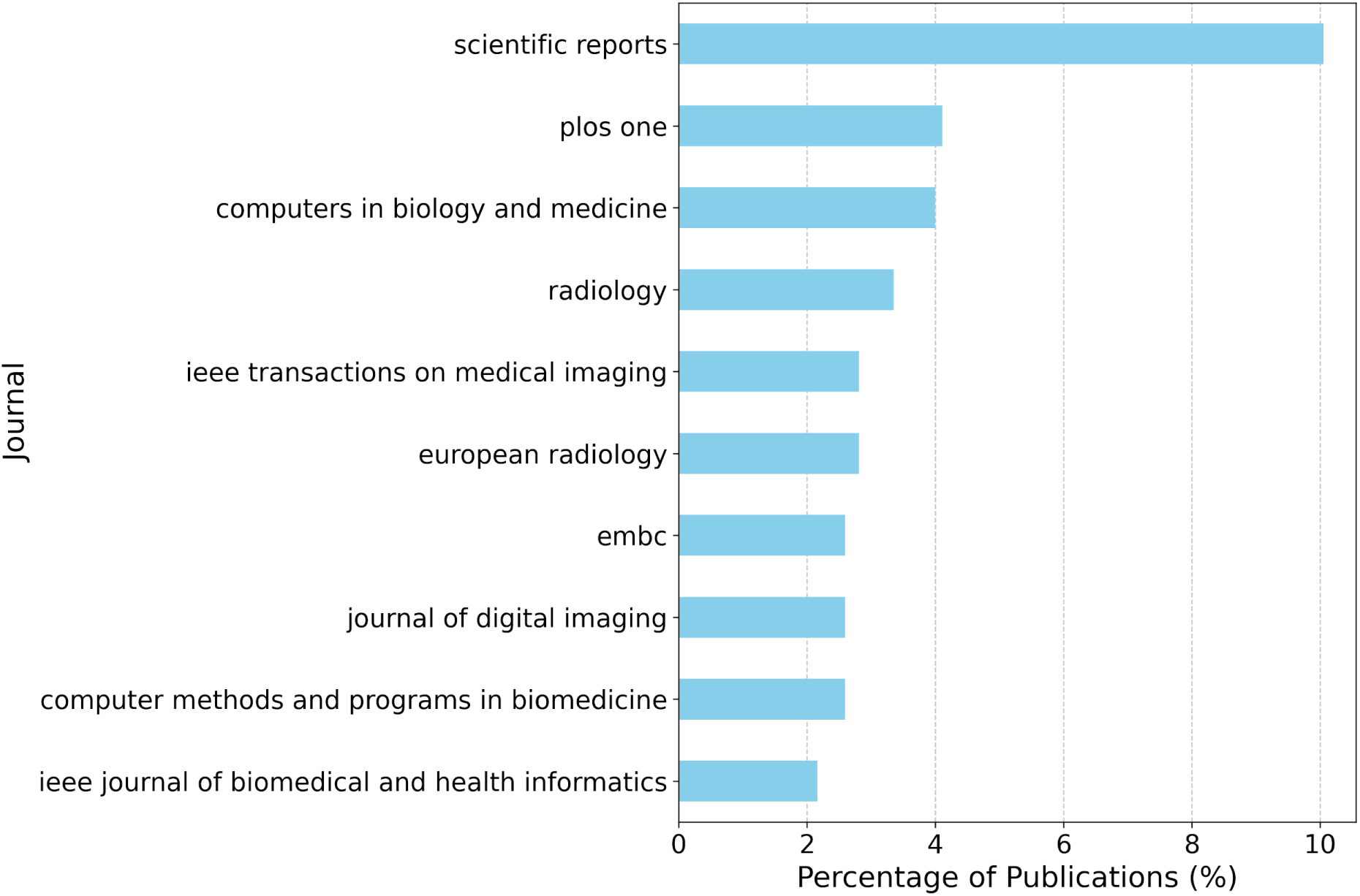
Journal distribution by percentage of analyzed publications.

Among the authors in the analyzed articles (Figure 3), China and the United States (US) stand out as the leaders in the number of publications in the first author position. China ranks first with 157 publications (16.93%), followed by the United States with 155 publications (16.72%). Together with South Korea (11.65%) and India (9.06%), these four countries account for more than half of the total publications. The distribution at the senior author position remains similar, with the United States at 18.00%, China at 15.84%, South Korea at 12.26%, and India at 7.81%.

**Fig. 3.**
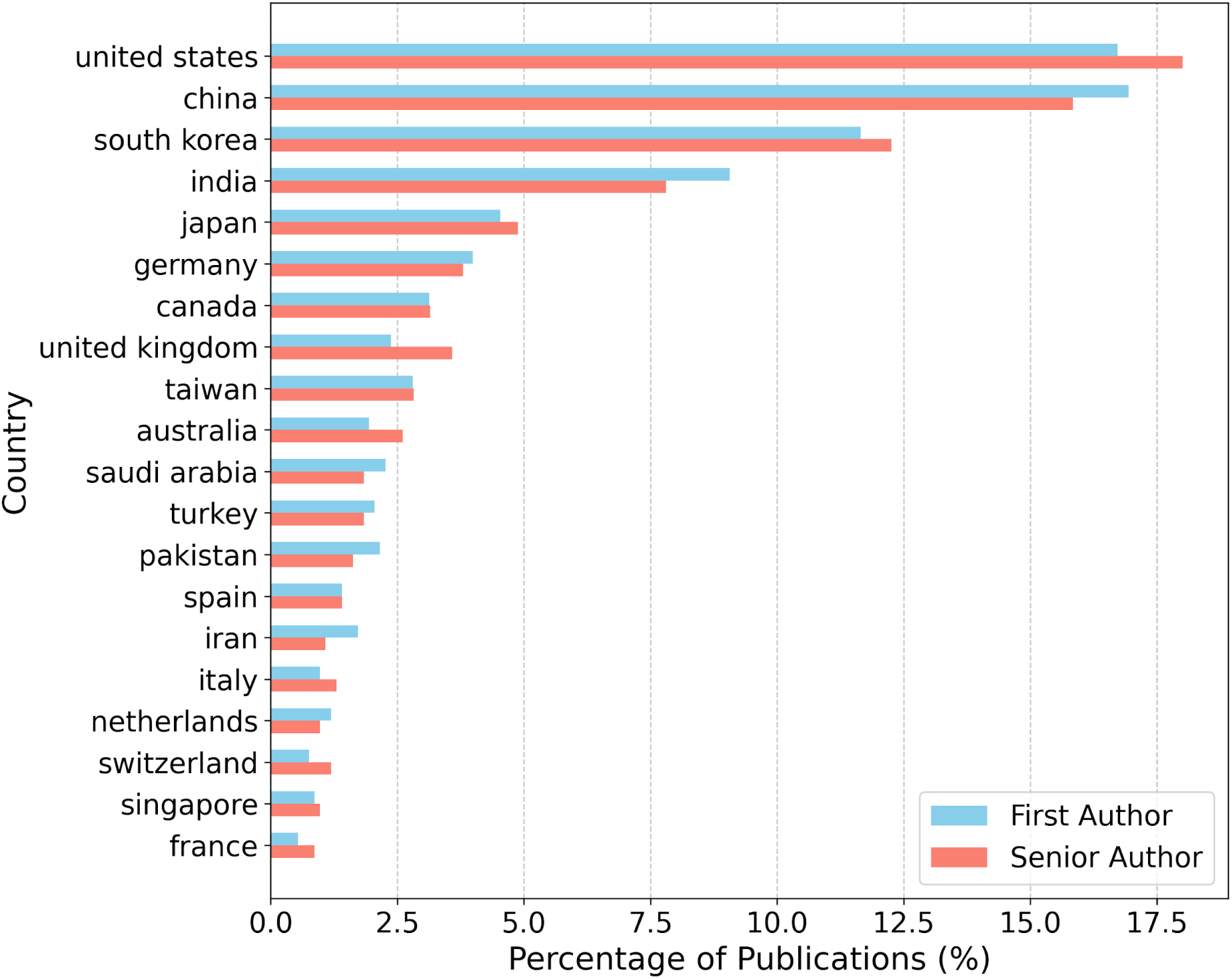
Distribution of articles in first and senior authors by country.

The ranking shifts when normalizing the author contributions by population, GDP, and R&D investment (Figure 4). Countries like Singapore, Qatar, Denmark, and Switzerland emerge as prominent contributors in per capita terms, while Jordan, Pakistan, and Tunisia, produce a relatively high volume of research when adjusted for GDP. Similarly, countries such as India, Pakistan, China, and Iraq show strong representation when adjusting for R&D investment.

**Fig. 4.**
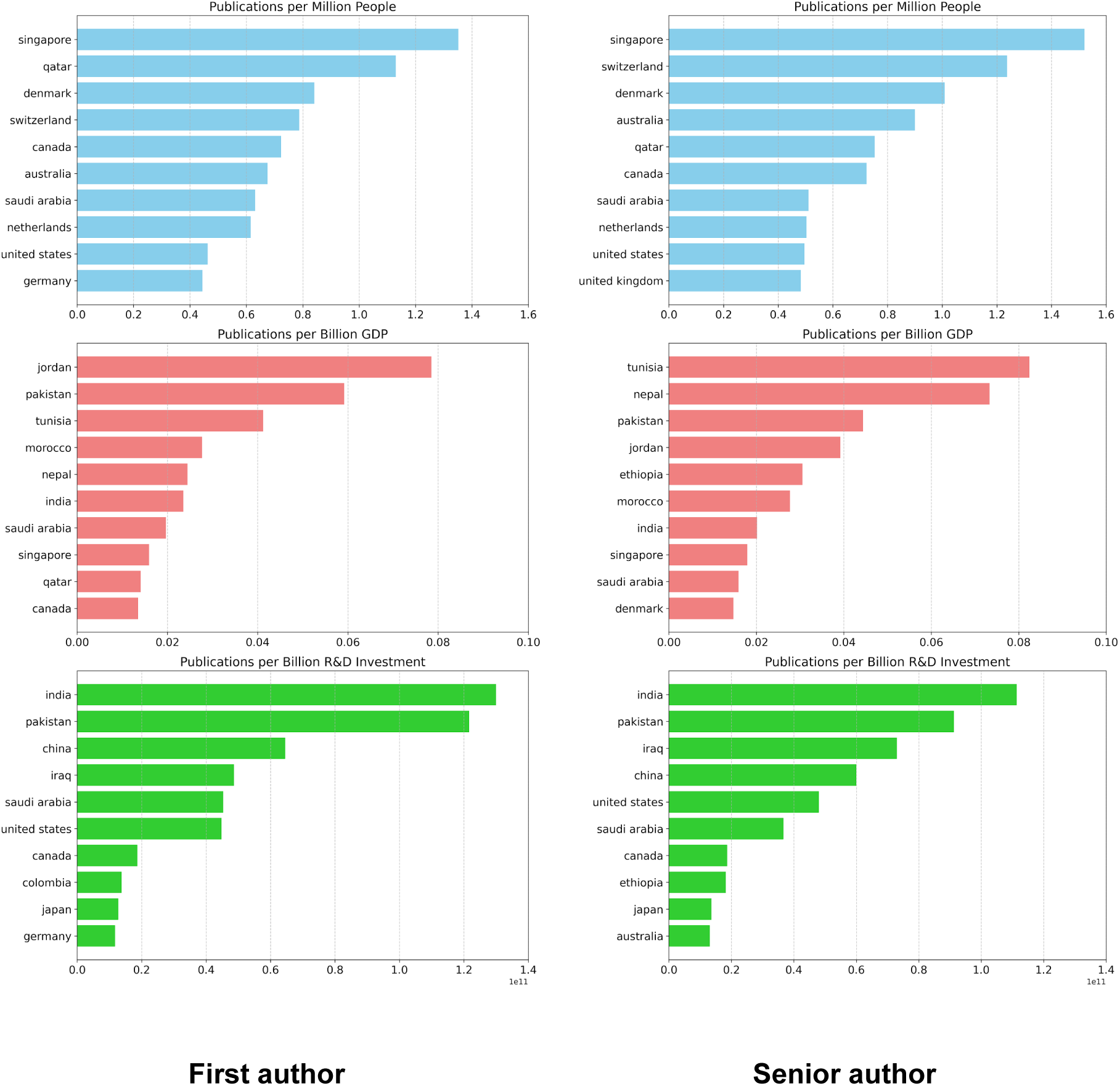
Distribution of publications adjusted for population size, GDP, and R&D investment, for first (left) and senior author (right).

An analysis of author income levels (Figure 5) shows that first authors were primarily affiliated with high-income countries (HICs, 55.6%), followed by upper-middle-income countries (UMICs, 25.8%) and lower-middle-income countries (LMICs, 18.6%). For senior authors, the trend was similar, with 59.7% from HICs, 24.2% from UMICs, and 15.4% from LMICs. Notably, no first authors were from low-income countries (LICs), and only 0.7% of senior authors had LIC affiliations.

**Fig. 5.**
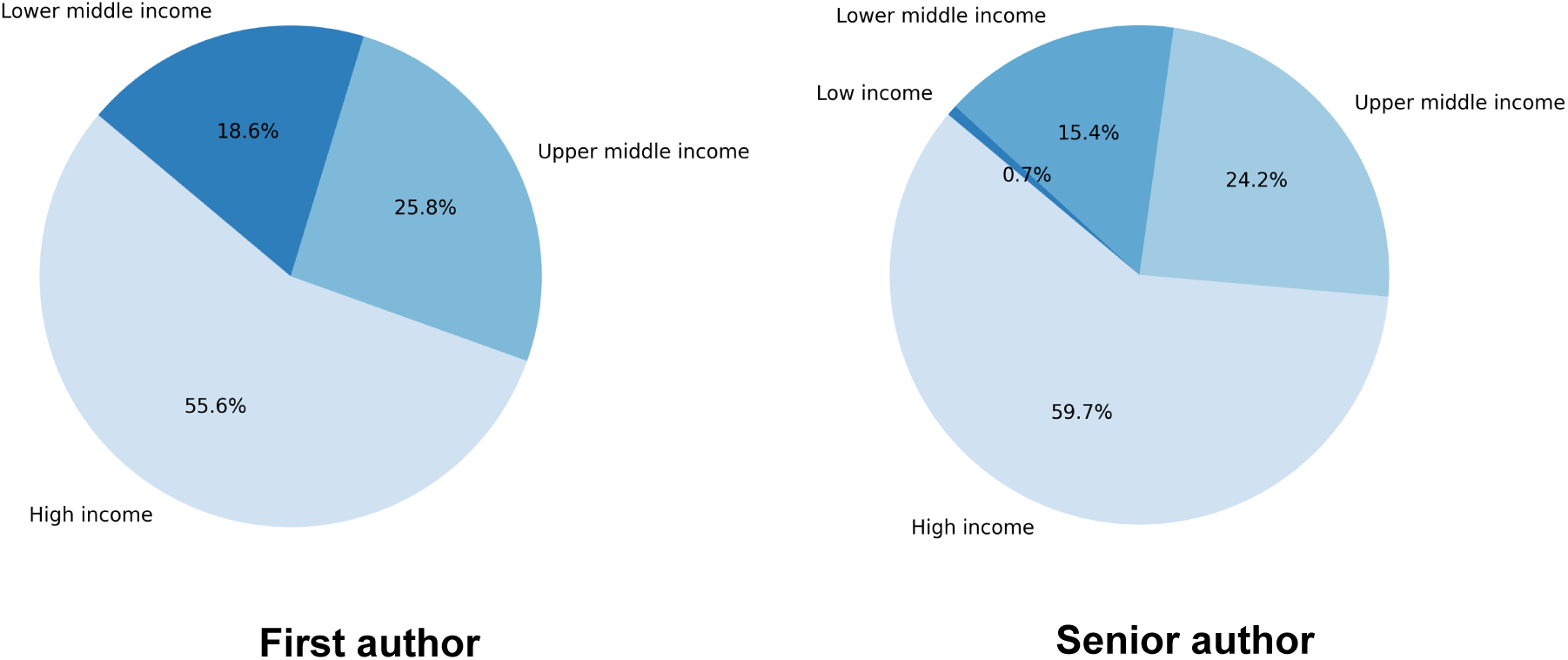
Income level representation of the first (left) and senior author (right).

No single institution demonstrated overwhelming dominance in either first or senior authorship, as even the leading contributors accounted for less than 3% of total publications. For first authorship, Seoul National University led (2.59%), followed by Seoul National University Hospital and the University of Toronto (1.19% each), and Yonsei University (1.08%).

A comparable distribution was observed for senior authorship, with Seoul National University remaining the most frequent affiliation (2.49%), followed by the University of Toronto (1.30%) and Seoul National University Hospital (1.19%). Harvard University and the University of Ulsan also emerged as notable contributors at the senior level (0.87% each).

Collaboration patterns (Figure 6), reveal a marked predominance of partnership between HIC institutions, with 53.43% among all analyzed articles. In contrast cross-income collaborations were comparatively rare. Partnerships with LMICs first authors and HIC senior authors (LMICs-HICs) accounted for 3.91% of publications, whereas HICs-LMICs collaboration represented only 1.68%.

**Fig. 6.**
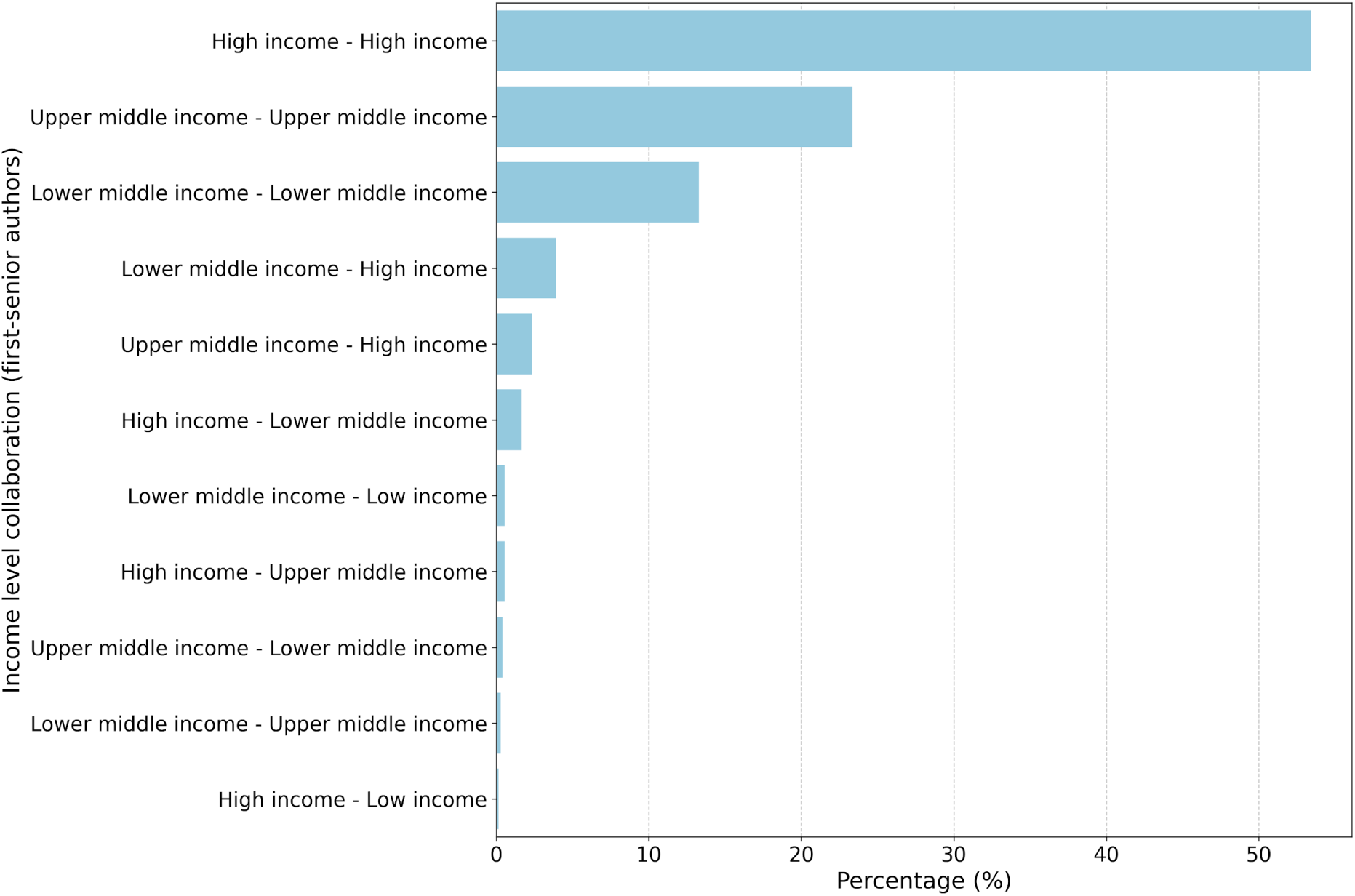
Collaboration patterns between first and senior authors.

Regarding dataset use (Figure 7), private datasets were the most common, accounting for 20.52% of all cases. Among public datasets, *ChestX-ray14* (8.24%) was the most frequently used, followed by *Chexpert (*6.31%) and the *COVID-19 Image Data Collection* (6.22%). The majority of datasets originated (Figure 8 and Figure 9) from HICs (73.6%), with the US being the largest single contributor (40.45%), followed by globally sourced datasets (20.35%), and China (14.80%).

**Fig. 7.**
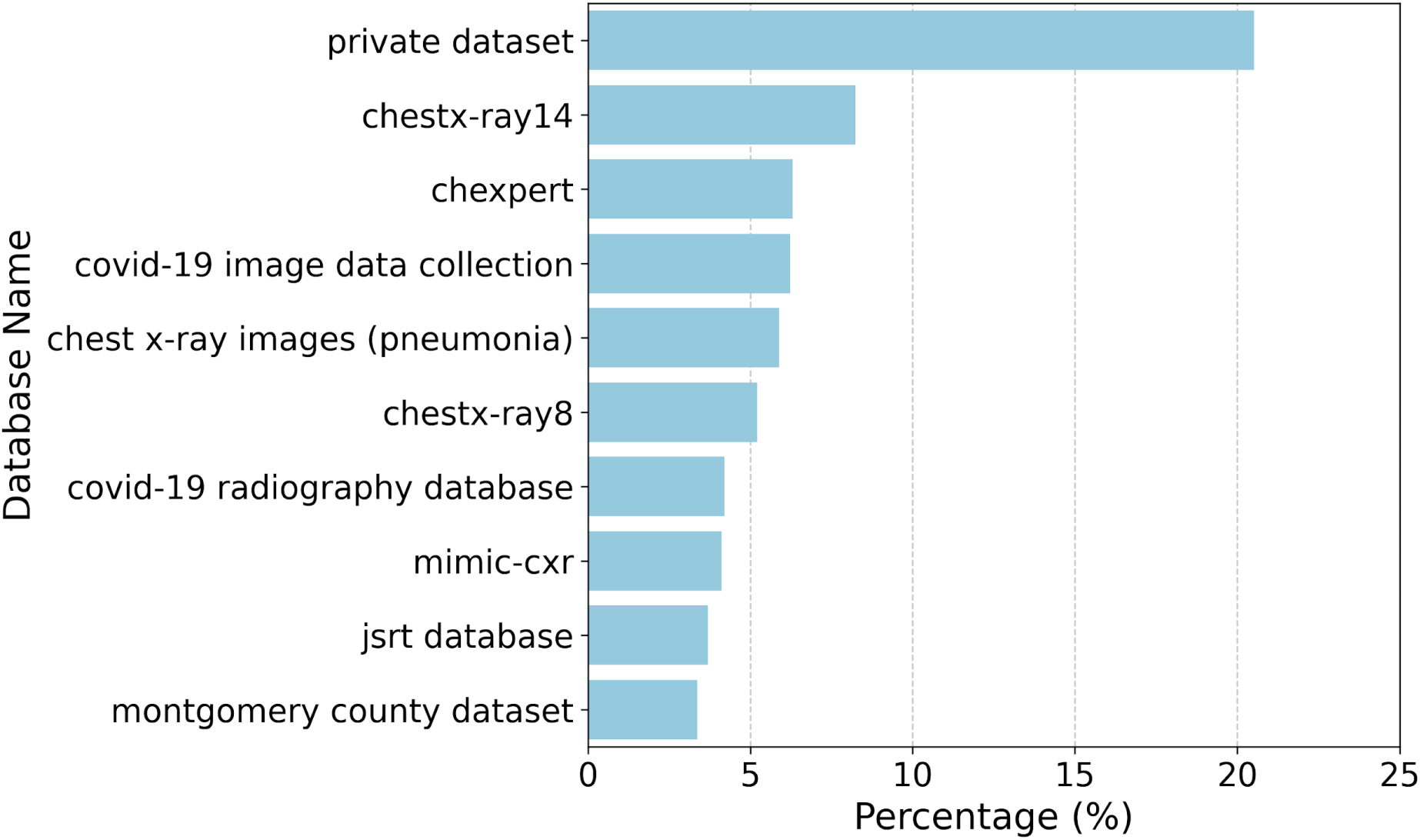
Most frequently used datasets in computer vision research for chest X-rays.

**Fig. 8.**
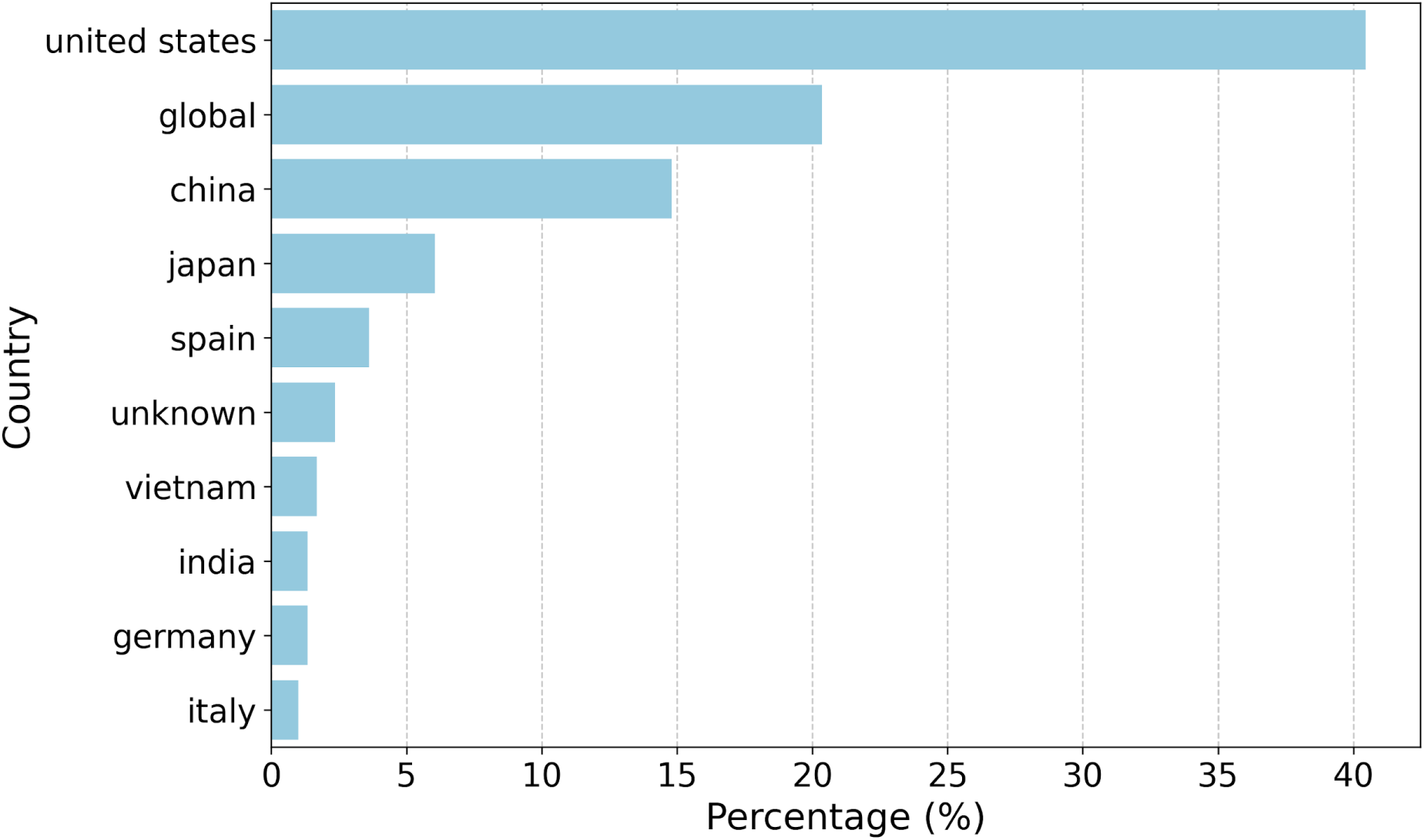
Country distribution of dataset sources.

**Fig. 9.**
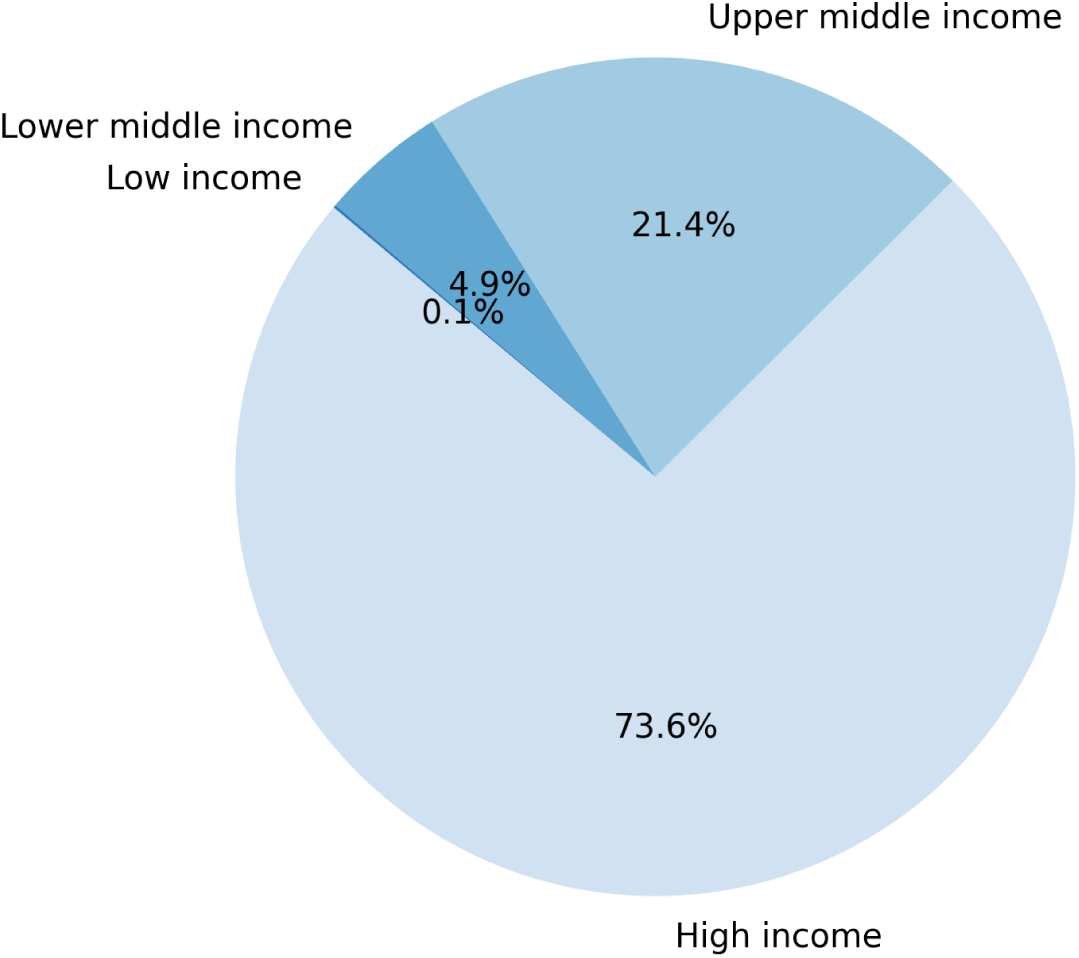
Income level distribution of dataset sources.

The relationship between dataset income level and author affiliation further highlights this concentration (Figure 10). Among studies using datasets from HICs, 51.38% had first authors affiliated with high-income institutions. In contrast, only 2.65% of publications involved datasets from LMICs with first authors from lower-middle-income institutions. A similar pattern was observed for senior authorship. Studies using HIC datasets most frequently had senior authors affiliated with high-income institutions (53.04%), whereas senior authors from lower-middle-income countries accounted for only 2.32% of publications involving lower-middle-income datasets.

**Fig. 10.**
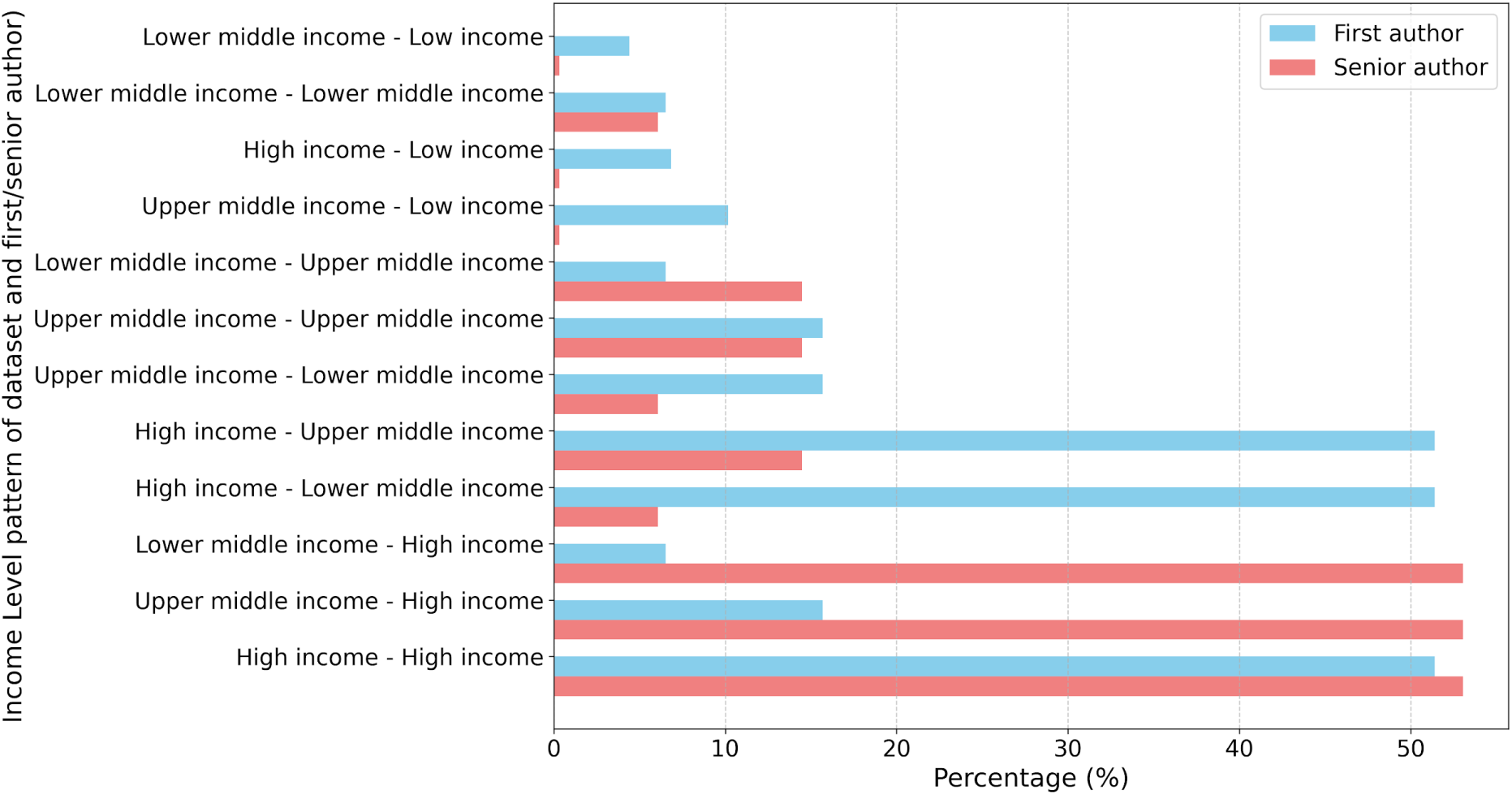
Relationship between dataset income level and author income level.

## Discussion

The results of this scientometric analysis reveal a landscape of pronounced global disparities in the field of computer vision for chest X-ray interpretation. The dominance of HICs in both authorship leadership and dataset provenance creates a self-reinforcing ecosystem that risks excluding the settings where AI-enabled clinical decision support is most urgently needed. These findings carry profound implications for the development of equitable, globally relevant medical AI technologies.

### 1. The leadership gap and its implications

We found that research leadership is heavily concentrated in HICs, which account for 55.6% of first authors and 59.7% of senior authors. This imbalance suggests that research priorities are predominantly shaped by high-resource perspectives, potentially overlooking the specific clinical needs for lower-middle-income countries where respiratory diseases carry a disproportionate burden.^3^ Notably, the total absence of first authors from low-income countries indicates that these populations have minimal voice in guiding the technologies intended for their healthcare systems. Representation in authorship is not merely a matter of prestige; it directly influences problem selection, the choice of evaluation metrics, and implementation strategies^19–24^.

### 2. Data poverty and the generalizability gap

The observed dataset provenance patterns further amplify these concerns. Our finding that 73.6% of training datasets originate from HICs, with the United States alone contributing 40.45%, suggests that most CV models are being developed on data that may not even reflect the diverse clinical presentations, comorbidity patterns, and radiographic characteristics of global populations. Previous studies have demonstrated that medical AI systems trained on demographically and geographically limited datasets exhibit significant performance deterioration when deployed in diverse settings.^25,26^ For instance, Yu et al. (2022) found that 81% of radiological deep learning algorithms experienced significant performance degradation when tested on external datasets, despite reporting high accuracy in internal validation.^27^ This issue is compounded by the preponderance of private datasets (20.52%), as closed data ecosystems limit transparency, external validation, and the iterative improvement of models across diverse populations.

### 3. Structural inequities and barriers to collaboration

The relationship between dataset provenance and author affiliation highlights a troubling barrier to data accessibility: over half of the studies using HIC datasets also featured HIC-affiliated lead authors. This mutually reinforcing relationship creates a scenario where technological insights and datasets flow predominantly within high-income circles without adequate bidirectional exchange. Such dynamics perpetuate power imbalances in knowledge production and lead to the implementation of technologies ill-suited to local contexts.^28,29^ Furthermore, structural inequities such as limited funding for imaging infrastructure and unequal access to computational resources further hamper meaningful participation from LMIC researchers.^30,31^ The fact that cross-income collaborations represent only 3.9% of publications suggests that current international partnerships are insufficient to bridge these gaps.

### 4. Roadmap for an equitable research ecosystem

Addressing these disparities requires coordinated action across multiple domains. First, funding agencies and global health organizations must prioritize developing diverse, well-curated, open-access CXR datasets from underrepresented populations. Second, research institutions should establish equitable international collaborations that engage LMIC partners as leaders rather than simply data sources, embedding principles of data sovereignty and shared governance. Third, publishers and conference organizers should adopt policies to incentivize diverse authorship and cross-income collaborations, potentially through dedicated submission tracks, mentorship programs, and reviewer training on geographic bias.

Researchers in high-income settings bear particular responsibility for challenging the status quo. This includes actively seeking collaborators from diverse geographic settings, sharing computational resources and methodological expertise, advocating for open-access publication models that do not disadvantage LMIC researchers, and challenging funding structures that limit international collaborations. Simultaneously, capacity-building initiatives within LMIC remain essential for sustainable change.

### 5. Conclusion

Our scientometric analysis reveals significant global disparities: high-income countries dominate authorship leadership (55.6% of first authors, 59.7% of senior authors) and dataset development (73.6% of sources). The near absence of authors from low-income countries, prevalence of private datasets (20.52%), and the rarity of cross-income collaborations (3.9%) suggest that AI diagnostic tools are being developed within a skewed ecosystem that may not adequately represent global healthcare needs.

These findings underscore the urgent need for structural reforms across the research landscape, ranging from the development of geographically diverse, open-access datasets to the establishment of equitable international partnerships that position LMIC researchers as knowledge producers. Only through deliberate efforts to democratize the production and application of medical AI we can ensure these technologies fulfill their promise of improving healthcare access and quality for all populations, regardless of geographic or economic circumstances.

## Materials and Methods

### I. Data Collection

This systematic review and scientometric analysis focused on the literature available for CV models applied to CXR. The study followed the PRISMA 2020 (Preferred Reporting Items for Systematic Reviews and Meta-Analyses) guidelines for reporting. The completed PRISMA 2020 checklist is provided in Supplementary Material 2. An automatic search was conducted across three databases: PubMed, Embase, and Scielo, using common terms related to CV and CXR. No language restrictions were applied. Refer to Supplementary Material 1 for the full search strategy.

The authors performed screening and eligibility assessment using the Covidence software, which automatically removed duplicate records. The article selection process followed two steps: (1) title and abstract screening, and (2) full-text eligibility assessment. Each step involved two independent reviews, with a third reviewer in case of disagreement.

Inclusion criteria comprised peer-reviewed journal and conference papers published between 2017 and 2025 that focused on CV or multimodal approaches applied to CXR. Preprints, review articles (surveys), incomplete papers, and studies not employing CV-based models (e.g., natural language processing or traditional machine learning methods) were excluded.

### II. Scientometric Analysis

To assess the research landscape of computer vision applied to chest X-rays, we performed a scientometric analysis of the 928 included studies. Using Dimensions AI API, we extracted information on publication journal, institutional affiliations, and authorship patterns, focusing on the institution and country of the first and last authors to evaluate global research contributions. To extract the information, we identified the Digital Object Identifiers (DOI) of all eligible studies and queried the Dimensions AI API. The retrieved data included authors’ names, institutional affiliations, and journal details, allowing us to determine the corresponding geographical distribution of research contributions. When a DOI was unavailable in the Dimensions database, we manually extracted the relevant information. Traditional clinical risk-of-bias tools were not applicable as this systematic review analyzed scientometric metadata rather than clinical outcomes; instead, we implemented a quality control process where two independent researchers manually verified institutional affiliations for missing DOIs, and country income levels were objectively classified using World Bank indicators to mitigate potential reporting biases.

After retrieving the countries, we classified them into income groups (low, lower-middle, upper-middle, and high-income) based on the World Bank world development indicators by income and region. Additionally, we manually extracted the databases referenced in each article and their country of origin.

To assess the relative contribution of each country, we normalized the number of publications using three factors: total population, gross domestic product (GDP), and Research and Development (R&D) investment. This normalization was performed to account for differences in country size and economic capacity, providing a more equitable comparison of research output. Population, GDP, and research investment data were obtained from the World Bank Open Data.

### III. Data Availability

The complete dataset of 928 analyzed publications and the extracted metadata used for this scientometric analysis will be made available as Supporting Information files upon publication. Requests for additional information regarding the analytical procedures can be directed to the corresponding author.

