## Supplementary Material 2 for "Who is leading medical AI? A systematic review and scientometric analysis of chest x-ray research"

**Supplementary Material 2 - Search Strategy**

1) **Pubmed**

| Search | PubMed Query | Item |
|---|---|---|
| #8 | #4 AND #7 | 1,248 |
| #7 | #5 AND #6 | 21,780 |
| #6 | (chest radiographs[Title/Abstract] OR chest radiography[Title/Abstract] OR chest imaging[Title/Abstract] OR chest X-ray*[Title/Abstract]) | 46,931 |
| #5 | (((Diagnostic Imaging[MeSH Terms]) OR (Imaging Diagnostic[Title/Abstract])) OR (Medical Imaging[Title/Abstract])) OR (Imaging Medical[Title/Abstract]) | 3,037,358 |
| #4 | #1 OR #2 OR #3 | 395,169 |
| #3 | ((((Deep Learning[MeSH Terms]) OR (Deep Learning[Title/Abstract])) OR (Learning Deep[Title/Abstract])) OR (Hierarchical Learning[Title/Abstract])) OR (Learning Hierarchical[Title/Abstract]) | 79,429 |
| #2 | ((((Machine Learning[MeSH Terms]) OR (Machine Learning[Title/Abstract])) OR (Learning Machine[Title/Abstract])) OR (Transfer Learning[Title/Abstract])) OR (Learning Transfer[Title/Abstract]) | 176,197 |
| #1 | (((((((((Artificial Intelligence[MeSH Terms]) OR (Artificial Intelligence[Title/Abstract])) OR (Computational Intelligence[Title/Abstract])) OR (Machine Intelligence[Title/Abstract])) OR (Computer Reasoning[Title/Abstract])) OR (AI[Title/Abstract])) OR (Computer Vision Systems[Title/Abstract])) OR (Computer Vision System[Title/Abstract])) OR (Knowledge Acquisition Computer[Title/Abstract])) OR (Knowledge Representation Computer[Title/Abstract])) OR (Knowledge Representations Computer[Title/Abstract]) | 298,995 |

((((((((((Artificial Intelligence[MeSH Terms]) OR (Artificial Intelligence[Title/Abstract])) OR (Computational Intelligence[Title/Abstract])) OR (Machine Intelligence[Title/Abstract])) OR (Computer Reasoning[Title/Abstract])) OR (AI[Title/Abstract])) OR (Computer Vision Systems[Title/Abstract])) OR (Computer Vision System[Title/Abstract])) OR (Knowledge Acquisition Computer[Title/Abstract])) OR (Knowledge Representation Computer[Title/Abstract])) OR (Knowledge Representations Computer[Title/Abstract]) OR ((((Machine Learning[MeSH Terms]) OR (Machine Learning[Title/Abstract])) OR (Learning Machine[Title/Abstract])) OR (Transfer Learning[Title/Abstract])) OR (Learning Transfer[Title/Abstract]) OR ((((Deep Learning[MeSH Terms]) OR (Deep Learning[Title/Abstract])) OR (Learning Deep[Title/Abstract])) OR (Hierarchical Learning[Title/Abstract])) OR (Learning Hierarchical[Title/Abstract])) AND ((((Diagnostic Imaging[MeSH Terms]) OR (Imaging Diagnostic[Title/Abstract])) OR (Medical Imaging[Title/Abstract])) OR (Imaging Medical[Title/Abstract]) AND (chest radiographs[Title/Abstract] OR chest radiography[Title/Abstract] OR chest imaging[Title/Abstract] OR chest X-ray*[Title/Abstract]))

(Artificial Intelligence OR Computational Intelligence OR Machine Intelligence OR Computer Reasoning OR AI OR Computer Vision Systems OR Computer Vision System OR Knowledge Acquisition Computer OR Knowledge Representation Computer OR Knowledge Representations Computer OR Machine Learning OR Machine Learning OR Learning Machine OR Transfer Learning OR Learning Transfer OR Deep Learning OR Deep Learning OR

Learning Deep OR Hierarchical Learning OR Learning Hierarchical) AND ((Diagnostic Imaging OR Imaging Diagnostic OR Medical Imaging OR Imaging Medical) AND (chest radiographs OR chest radiography OR chest imaging OR chest X-ray*))

### 2) Embase

| Search | The Embase | Item |
|---|---|---|
| #16 | #10 AND #15 | 667 |
| #15 | #13 AND #14 | 3,697 |
| #14 | 'chest radiographs':ab,kw,ti OR 'chest radiography':ab,kw,ti OR 'chest X-ray*':ab,kw,ti OR 'chest imaging':ab,kw,ti | 86,714 |
| #13 | #11 OR #12 | 321,901 |
| #12 | 'diagnostic imaging':ab,kw,ti OR 'imaging diagnostic':ab,kw,ti OR 'medical imaging':ab,kw,ti OR 'imaging medical':ab,kw,ti | 56,571 |
| #11 | 'diagnostic imaging'/exp | 294,163 |
| #10 | #3 OR #6 OR #9 | 750,147 |
| #9 | #7 OR #8 | 107,585 |
| #8 | 'deep learning':ab,kw,ti OR 'learning deep':ab,kw,ti OR 'hierarchical learning':ab,kw,ti OR 'learning hierarchical':ab,kw,ti | 97,973 |
| #7 | 'deep learning'/exp | 81,601 |
| #6 | #4 OR #5 | 614,717 |
| #5 | 'machine learning':ab,kw,ti OR 'learning machine':ab,kw,ti OR 'transfer learning':ab,kw,ti OR 'learning transfer':ab,kw,ti | 180,829 |
| #4 | 'machine learning'/exp | 592,861 |
| #3 | #1 OR #2 | 225,464 |
| #2 | 'artificial intelligence':ab,kw,ti OR 'computational intelligence':ab,kw,ti OR 'machine intelligence':ab,kw,ti OR 'computer reasoning':ab,kw,ti OR 'ai':ab,kw,ti OR 'computer vision systems':ab,kw,ti OR 'computer vision system':ab,kw,ti OR 'knowledge acquisition computer':ab,kw,ti OR 'knowledge representation computer':ab,kw,ti OR 'knowledge representations computer':ab,kw,ti | 162,590 |
| #1 | 'artificial intelligence'/exp | 143,062 |
